# Convalescent Plasma for COVID-19. A randomized clinical trial

**DOI:** 10.1101/2020.07.01.20139857

**Authors:** Arvind Gharbharan, Carlijn C.E. Jordans, Corinne Geurtsvankessel, Jan G. den Hollander, Faiz Karim, Femke P. N. Mollema, Janneke E. Stalenhoef-Schukken, Anthonius Dofferhoff, Inge Ludwig, Adrianus Koster, Robert-Jan Hassing, Jeannet C. Bos, Geert R. van Pottelberge, Imro N. Vlasveld, Heidi S. M. Ammerlaan, Elena M. van Leeuwen-Segarceanu, Jelle Miedema, Menno van der Eerden, Grigorios Papageorgiou, Peter te Boekhorst, Francis H. Swaneveld, Peter D. Katsikis, Yvonne Mueller, Nisreen M.A. Okba, Marion P.G. Koopmans, Bart H.G. Haagmans, Casper Rokx, Bart J.A. Rijnders

## Abstract

**Background:** After recovery from COVID-19, most patients have anti-SARS-CoV-2 neutralizing antibodies. Their convalescent plasma could be an inexpensive and widely available treatment for COVID-19.

**Methods:** The Convalescent-plasma-for-COVID (ConCOVID) study was a randomized trial comparing convalescent plasma with standard of care therapy in patients hospitalized for COVID-19 in the Netherlands. Patients were randomized 1:1 and received 300ml of plasma with anti-SARS-CoV-2 neutralizing antibody titers of at least 1:80. The primary endpoint was day-60 mortality and key secondary endpoints were hospital stay and WHO 8-point disease severity scale improvement on day 15.

**Results:** The trial was halted prematurely after 86 patients were enrolled. Although symptomatic for only 10 days (IQR 6-15) at the time of inclusion, 53 of 66 patients tested had anti-SARS-CoV-2 antibodies at baseline. A SARS-CoV-2 plaque reduction neutralization test showed neutralizing antibodies in 44 of the 56 (79%) patients tested with median titers comparable to the 115 donors (1:160 vs 1:160, p=0.40). These observations caused concerns about the potential benefit of convalescent plasma in the study population and after discussion with the data safety monitoring board, the study was discontinued. No difference in mortality (p=0.95), hospital stay (p=0.68) or day-15 disease severity (p=0.58) was observed between plasma treated patients and patients on standard of care.

**Conclusion:** Most COVID-19 patients already have high neutralizing antibody titers at hospital admission. Screening for antibodies and prioritizing convalescent plasma to risk groups with recent symptom onset will be key to identify patients that may benefit from convalescent plasma. Clinicaltrials.gov: NCT04342182

## Introduction

Severe acute respiratory syndrome coronavirus 2 (SARS-CoV-2) puts a tremendous strain on healthcare systems. A recently completed clinical trial demonstrated that *anti-inflammatory* therapy with dexamethasone significantly decreases overall mortality.^1^ Antivirals as lopinavir-ritonavir failed to show survival benefit ^2^, but a more recent randomized trial showed a shortened time to clinical recovery in patients treated with remdesivir and comparable results were observed in a trial on interferon beta-1b, lopinavir–ritonavir, and ribavirin. However, it remains to be seen if any of these antiviral therapies will decrease mortality.^3,4^ Also, these drugs are not widely available and the rapid distribution to hospitals across the world is extremely challenging.^1^ Therefore, other readily available, affordable and effective antiviral therapies are needed.

Convalescent plasma (ConvP), which may contain high levels of virus neutralizing antibodies, could be an alternative treatment option for SARS-CoV-2 patients. A similar strategy has been pursued during the 2003 SARS and later MERS outbreaks. ^5^ Conclusive evidence for the effectivity of ConvP as a treatment for human coronavirus infections has yet not been documented in large randomized clinical trials. Preclinical research however indicated a protective effect of human ConvP when given to hamsters infected with SARS-CoV-2 early in the disease course. ^6^ Although large volumes of ConvP can have indirect effects as well, we assume that key to the efficacy of ConvP through direct antiviral effect may be the presence of high titers of virus neutralizing antibodies. Following this rationale, benefit can only be expected if it is administered to viremic patients with little or no autologous neutralizing antibodies. Although ConvP seems to be safe, no overall clinical benefit of ConvP therapy was observed in a prematurely interrupted randomized trial from China.^7,8^ In this study, patients had been symptomatic for 30 days on average when they received ConvP which contrasts sharply with the time from symptom onset to hospital admission in cohort studies from China (11 days) and Europe (13 days).^9,10^ The recent observations showing that close to 100% of patients have detectable neutralizing antibodies three weeks after symptom onset may explain the lack of a therapeutic effect observed.^11,12^ All other evidence on the possible efficacy of ConvP comes from uncontrolled case series or with historical patients as comparator. ^13-15^ Despite the fact that in the USA already 8932 patients had received ConvP as part of the early access program on the 11^th^ of May 2020, it remains unknown if ConvP will be beneficial if administered early on in the COVID-19 disease course.^8^ The ConCOVID study was setup in centers across the Netherlands in order to determine the effect of ConvP on mortality in COVID-19 patients early after hospital admission.

## Methods

### Study design

The ConCOVID study was designed as a nationwide multicenter open-label randomized clinical trial. The trial network includes 14 secondary and academic hospitals in the Netherlands. Enrollment began on April 8 2020. Eligible patients were at least 18 years, admitted to a study site for COVID-19 and had clinical COVID-19 disease proven by a positive SARS-CoV-2 reverse transcriptase polymerase chain reaction (RT-PCR) test in the previous 96 hours. Patients with a documented IgA deficiency or on mechanical ventilation for >96 hours were excluded. ConvP donors were recruited and screened by Sanquin Blood Supply (the Dutch blood bank) according to existing guidelines. They needed to have had a RT-PCR confirmed SARS-CoV-2 infection and be asymptomatic for at least 14 days. Of all donors tested, only plasma with anti-SARS-CoV-2 neutralizing antibodies confirmed by a SARS-COV-2 plaque reduction neutralization test (PRNT) and a PRNT50 titer of at least 1:80 was used.^11^ Furthermore, for each patient, we selected the plasma with the highest PRNT50 titer from the ABO compatible donor pool available at the time of inclusion. Donors were asked to complete a detailed questionnaire on their medical history, COVID-19 related clinical symptoms and the symptom duration.

### Intervention, primary and secondary endpoints

Patients were randomly assigned via a web-based system at a 1:1 ratio to the current standard of care at each hospital with or without the addition of 300ml of ConvP, the standard volume of one plasma unit produced by Sanquin Blood Supply, was administered intravenously on the day of inclusion. Patients without a clinical response and a persistently positive RT-PCR could receive a second plasma unit after five days. Off-label use of EMA-approved drugs (e.g. chloroquine, azithromycin, lopinavir/ritonavir, tocilizumab, anakinra) as a treatment for COVID-19 was allowed in hospitals were this was part of the standard of care. We scored the clinical status with the ordinal 8-point WHO COVID-19 disease severity scale on days 1, 15 and 30.^16^ Serum samples and nasopharyngeal swabs were collected at inclusion preceding treatment and thereafter. The primary endpoint of the study was overall mortality until discharge from the hospital or a maximum of 60 days after admission whichever came first. Key secondary clinical endpoint we describe here are the improvement on the 8-point WHO COVID-19 disease severity scale from inclusion to day 15, hospital length of stay and safety.

### Assays

We analyzed serum samples of donors and patients for the presence of neutralizing antibodies by performing a PRNT with the SARS-CoV-2 virus (German isolate; GISAID ID EPI_ISL 406862; European Virus Archive Global #026V-03883) as we have described previously.^11^ More details are available in the online supplement. Serum was also tested for the presence of anti-SARS-CoV-2 total Ig and IgM with the Wantai Enzyme Linked Immunosorbent Essay (ELISA) test (Wantai Biological, Beijing). We previously showed that a positive total Ig or a IgM with an optical density (OD) ratio >10 (which equals an OD of 2.0), correlates closely with virus neutralizing antibody titers (PRNT50) of at least 1:80.^17^

### Sample size and statistical analysis plan

With an anticipated 50% overall mortality reduction from 20% in the control arm, which was the reported mortality in hospitalized patients in the Netherlands when the protocol was designed and with a control to intervention ratio of 1:1, 426 patients were needed for the study to have 80% power with a global alpha level of 0.05 and adjusted alpha level for the primary endpoint of 0.0480, accounting for 1 interim analysis. The full statistical analysis plan is available in the online supplement and the full protocol. Due to the premature interruption of the trial and resulting lower event rates we present both the results of the multivariable (adjusted) logistic regression analysis as originally planned and the unadjusted analysis (table 2 and 3 of online supplement).

The study was reviewed and approved by the institutional review board of the Erasmus University Medical Center. Written informed consent was obtained from every patient or a legal patient representative. The DSMB reviewed the safety of the participants on a regular basis and recommended the study team regarding the further conduct of the study at predefined time points. The study was registered as NCT04342182 at clinicaltrials.gov.

## Results

### Baseline characteristics of donors and patients

Of over 3200 ex-COVID-19 patients who volunteered to be screened for ConvP donation, the first 115 who fulfilled the COVID-19 donor criteria were screened for the presence of neutralizing antibodies. 100 of them also completed the questionnaire. Baseline characteristics are given in table 1. Most donor volunteers were males and part of the women were rejected as donor because of HLA/HNA antibodies. Donors had been symptomatic for a median of 12 days (IQR 8 - 18) and were younger compared to patients with a substantially milder disease course. Ninety-nine percent of the donors tested positive with the Wantai total Ig ELISA and neutralizing antibodies were detectable in 96% at a median PRNT50 titer of 1:160 (IQR 1:80 – 1:640); 78% and 43% had PRNT50 titers of at least 1:80 or 1:320 respectively (figure 1a). As defined in the protocol, for each newly enrolled patient we always used a plasma unit with the highest available PRNT50 titer. This resulted in the use of plasma from 19 donors and with a median titer of 1:640 (IQR 1:320 - 1:1280).

**Table 1.**
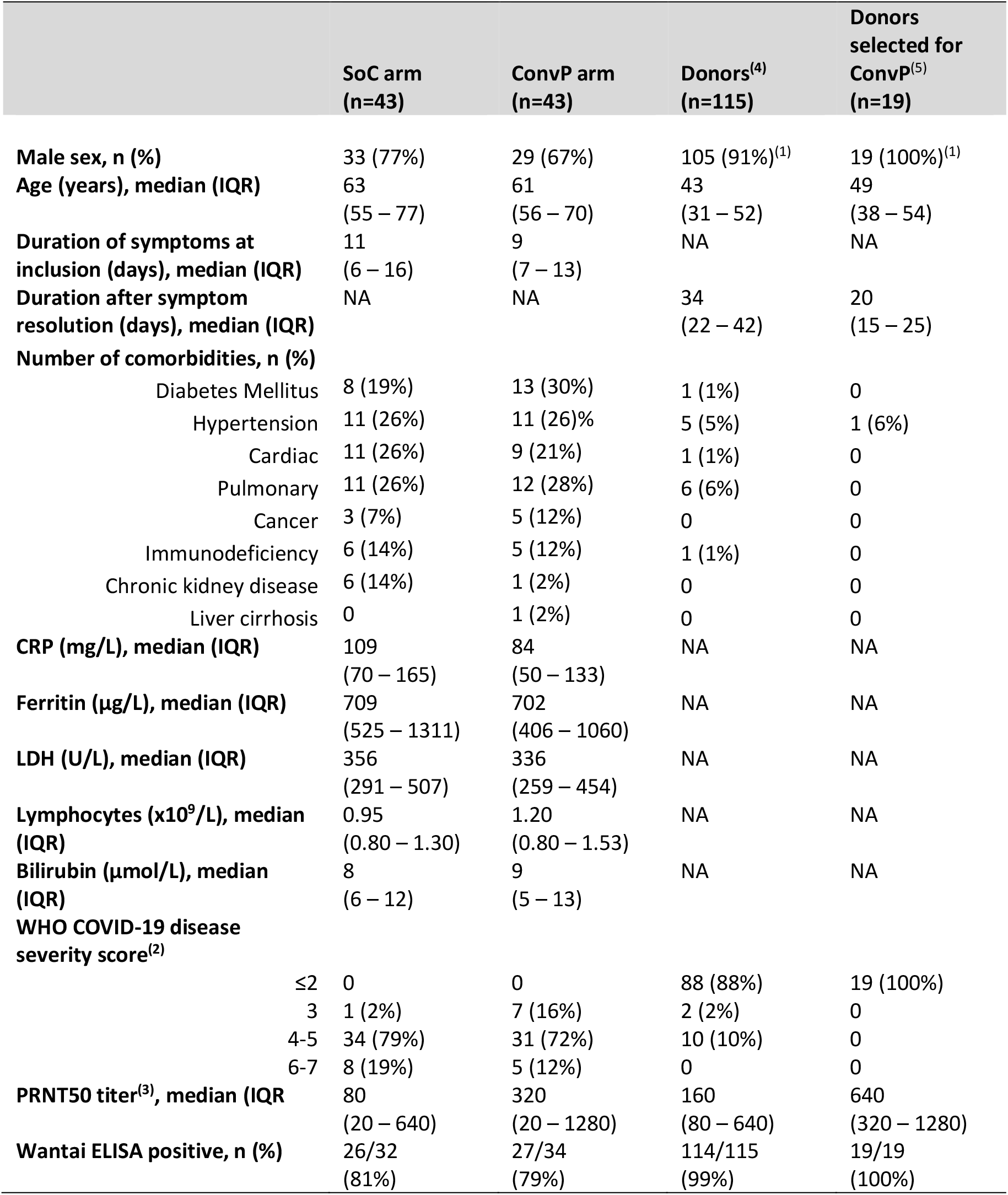
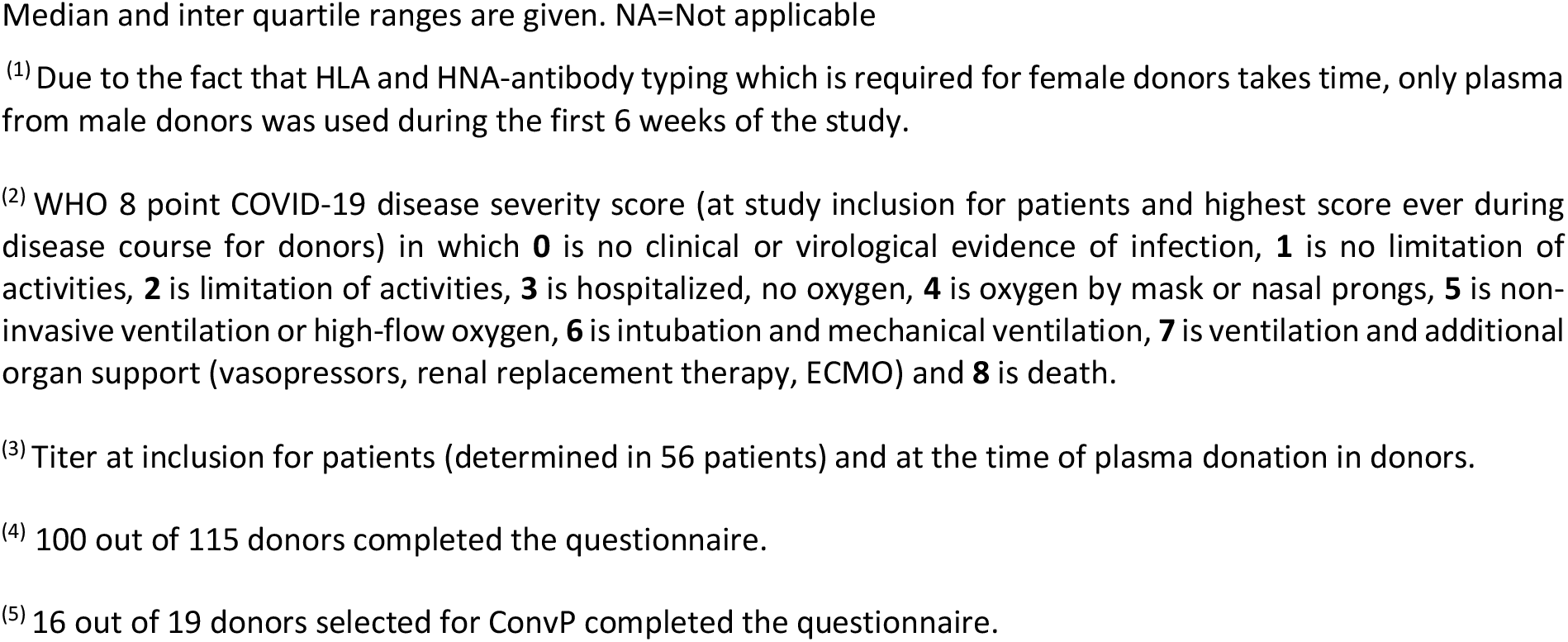
Baseline characteristics of patients and ConvP donors. Median and inter quartile ranges are given

|  | **SoC arm**<br>**(n=43)** | **ConvP arm**<br>**(n=43)** | **Donors<sup>(4)</sup>**<br>**(n=115)** | **Donors**<br>**selected for**<br>**ConvP<sup>(5)</sup>**<br>**(n=19)** |
| --- | --- | --- | --- | --- |
| **Male sex, n (%)** | 33 (77%) | 29 (67%) | 105 (91%)<sup>(1)</sup> | 19 (100%)<sup>(1)</sup> |
| **Age (years), median (IQR)** | 63<br>(55 – 77) | 61<br>(56 – 70) | 43<br>(31 – 52) | 49<br>(38 – 54) |
| **Duration of symptoms at**<br>**inclusion (days), median (IQR)** | 11<br>(6 – 16) | 9<br>(7 – 13) | NA | NA |
| **Duration after symptom**<br>**resolution (days), median (IQR)** | NA | NA | 34<br>(22 – 42) | 20<br>(15 – 25) |
| **Number of comorbidities, n (%)** |  |  |  |  |
| Diabetes Mellitus | 8 (19%) | 13 (30%) | 1 (1%) | 0 |
| Hypertension | 11 (26%) | 11 (26%) | 5 (5%) | 1 (6%) |
| Cardiac | 11 (26%) | 9 (21%) | 1 (1%) | 0 |
| Pulmonary | 11 (26%) | 12 (28%) | 6 (6%) | 0 |
| Cancer | 3 (7%) | 5 (12%) | 0 | 0 |
| Immunodeficiency | 6 (14%) | 5 (12%) | 1 (1%) | 0 |
| Chronic kidney disease | 6 (14%) | 1 (2%) | 0 | 0 |
| Liver cirrhosis | 0 | 1 (2%) | 0 | 0 |
| **CRP (mg/L), median (IQR)** | 109<br>(70 – 165) | 84<br>(50 – 133) | NA | NA |
| **Ferritin (µg/L), median (IQR)** | 709<br>(525 – 1311) | 702<br>(406 – 1060) | NA | NA |
| **LDH (U/L), median (IQR)** | 356<br>(291 – 507) | 336<br>(259 – 454) | NA | NA |
| **Lymphocytes (x10<sup>9</sup>/L), median**<br>**(IQR)** | 0.95<br>(0.80 – 1.30) | 1.20<br>(0.80 – 1.53) | NA | NA |
| **Bilirubin (µmol/L), median**<br>**(IQR)** | 8<br>(6 – 12) | 9<br>(5 – 13) | NA | NA |
| **WHO COVID-19 disease**<br>**severity score<sup>(2)</sup>** |  |  |  |  |
| ≤2 | 0 | 0 | 88 (88%) | 19 (100%) |
| 3 | 1 (2%) | 7 (16%) | 2 (2%) | 0 |
| 4-5 | 34 (79%) | 31 (72%) | 10 (10%) | 0 |
| 6-7 | 8 (19%) | 5 (12%) | 0 | 0 |
| **PRNT50 titer<sup>(3)</sup>, median (IQR)** | 80<br>(20 – 640) | 320<br>(20 – 1280) | 160<br>(80 – 640) | 640<br>(320 – 1280) |
| **Wantai ELISA positive, n (%)** | 26/32<br>(81%) | 27/34<br>(79%) | 114/115<br>(99%) | 19/19<br>(100%) |

**Figure 1a.**
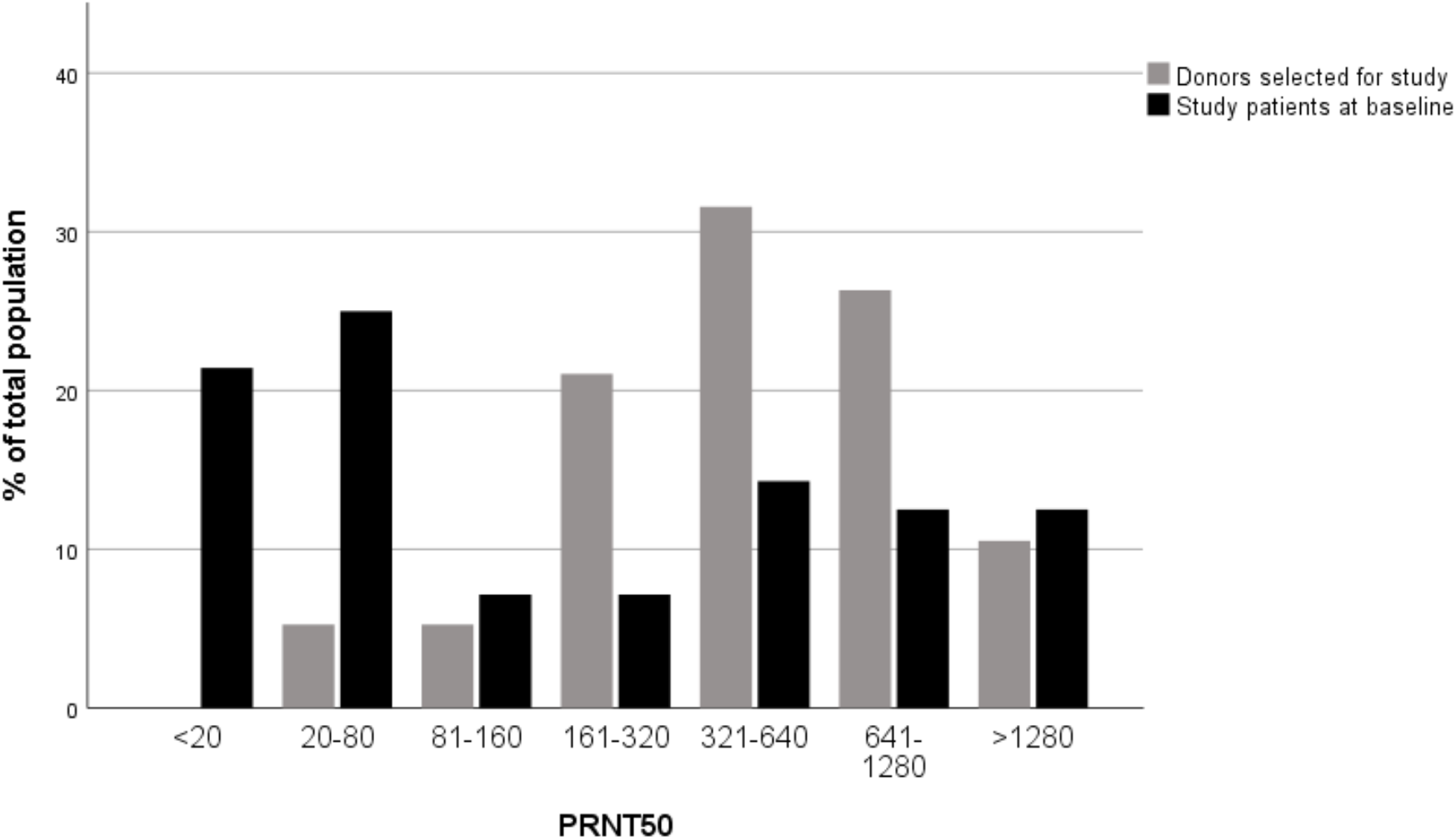
PRNT50 in - donors of whom ConvP was used in the study (grey) and in - study patients at baseline (black).

Of the 204 patients diagnosed with COVID-19 and evaluated for eligibility, 86 were enrolled when the study was halted. The most frequent reason for non-eligibility was informed consent refusal, typically due to fear of adverse events (figure 2). Patients were mostly males of 63 (IQR 56 – 74) years of age. At the time of inclusion, they had COVID-19 related symptoms for 10 days (IQR 6 – 15) and had been admitted to the hospital for 2 days (IQR 1 – 3 days). Thirteen patients were admitted directly to ICU and were on mechanical ventilation at inclusion (table 1). Blood samples of 66 patients could be collected. On the day of inclusion, 53 (80%) tested positive for anti-SARS-CoV-2 antibodies. The total Ig ratio was >10 in 39 (60%). The SARS-CoV-2 PRNT50 titer could be measured in 56 of these 66 and in 44 (79%) neutralizing antibodies at a titer of ≥1:20 were detected. The median PRNT50 titer in these 56 patients was comparable to the titer observed in the overall donor population (1:160 vs 1:160, p=0.40). The median PRNT50 titers of the plasma units actually used in the study were higher than titers of patients at inclusion (1:640 vs 1:160, p=0.01), table 1. Figure 1a and b illustrate the distribution of baseline PRNT50 levels in patients, the overall donor population and the 19 donors from whom ConvP was used.

**Figure 1b.**
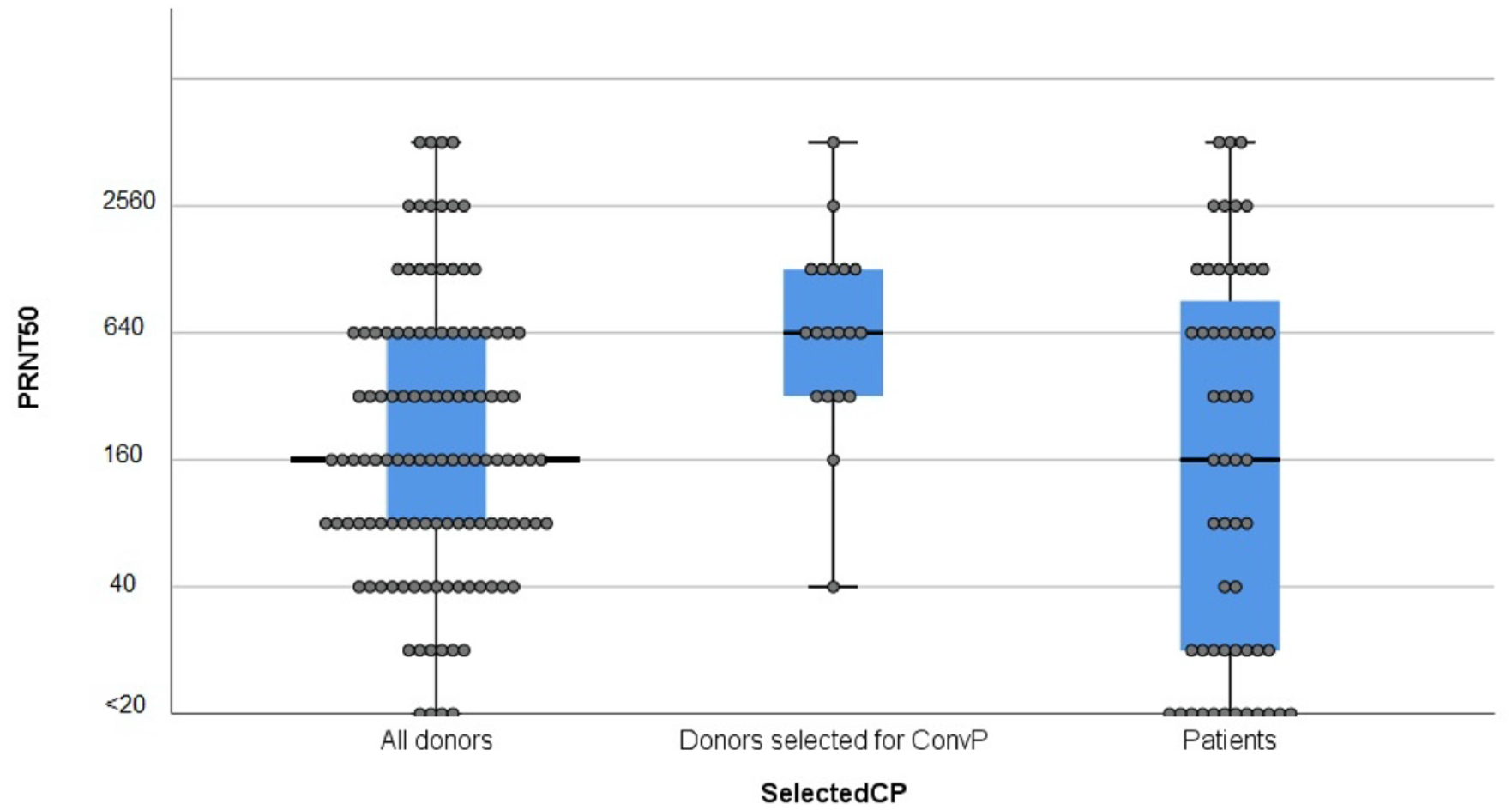
PRNT50 titers in overall donor population (n=115), donors of whom ConvP was used (n=19) and patients at baseline (n=56). P=0.398 for the difference between all donors and patients and p=0.011 for difference between Donors selected for ConvP and patients.

**Figure 2.**
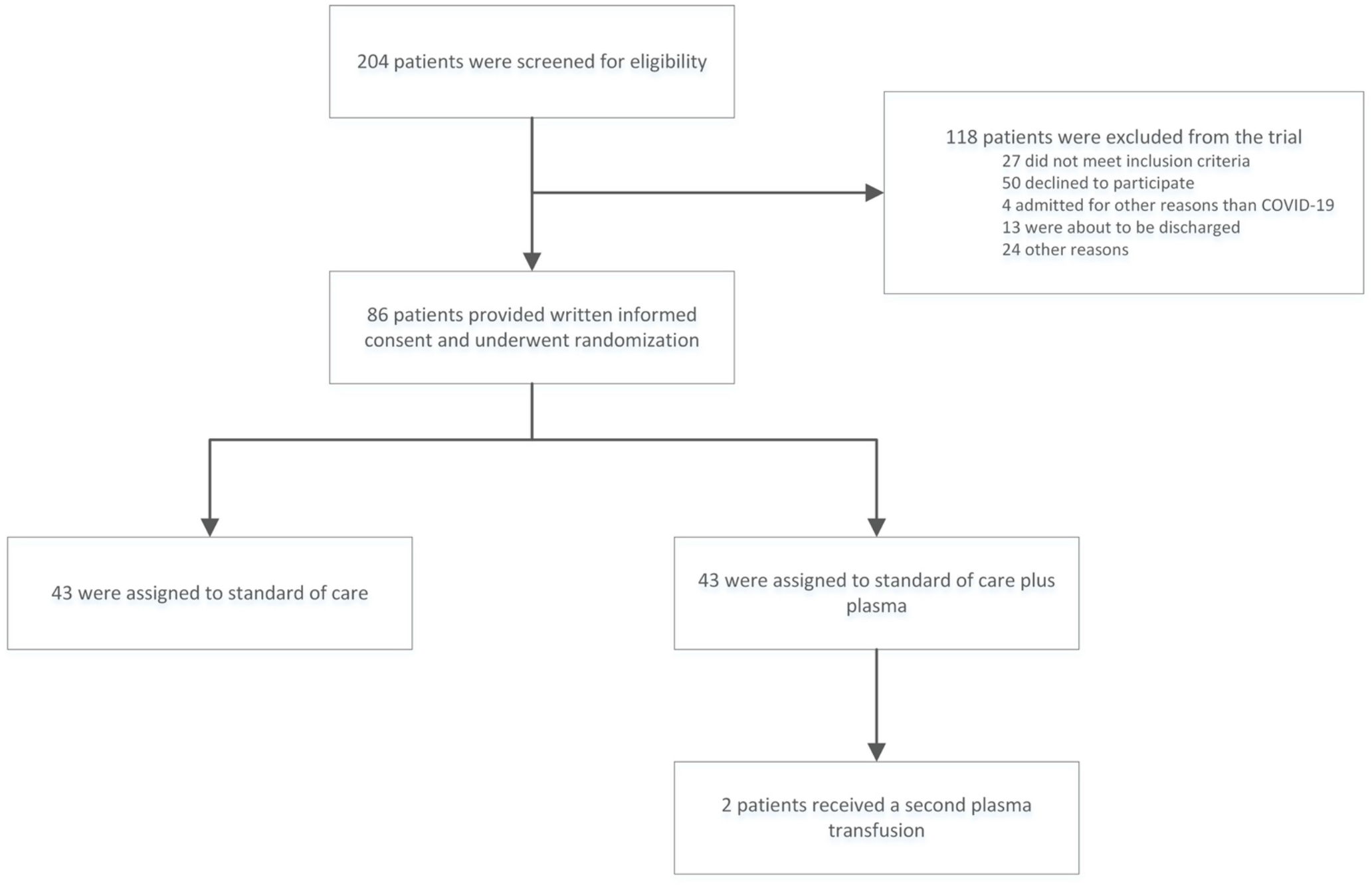
Patient flow in the study

To confirm the observation that the majority of patients already had high neutralizing antibody titers at hospital admission, we tested an additional 37 serum samples that had been collected within three days after hospital admission from COVID-19 patients admitted on a general ward at Erasmus MC in the weeks preceding the start of the ConCOVID study. With a median age of 65 years (IQR 56 – 74), 60% males and symptom duration of 9 days (IQR 4 - 13) these patients were comparable to the study population. Here as well we found that 26/37 (70%) of patients had anti-SARS-CoV-2 Ig antibodies and in 23/37 (62%) at a ratio >10, indicating high neutralization capacity.

In a post-hoc analysis we evaluated if ConvP administration accelerated the increase in neutralizing antibody titers over time. For this, the PRNT50 titers on day 7 were compared with the titers on day 1 in the subgroup of patients with a titer <1:160 at baseline. In all 9 patients, including 5 randomized to standard of care, a fourfold increase in titers was observed on day 7 (supplemental figure 1c).

### Efficacy and safety

The adjusted OR for overall mortality for patients treated with ConvP was 0.95 (CI 0.20 – 4.67., p=0.95). Of the 43 patients randomized to ConvP 6 (14%) had died while 11 of the 43 (26%) control patients had died. At that time, all 86 patients had been followed for at least 15 days after inclusion and 75 and 32 for at least 30 and 60 days respectively.

The adjusted OR for an improvement in the WHO COVID-19 disease severity score on day 15 was 1.30 (CI 0.52 - 3.32). Twenty-five (58%) of the patients in the plasma group and 25 (58%) in the control group showed an improvement in their WHO COVID-19 disease severity score on that day. Treatment with ConvP was not associated with a shorter time to discharge from the hospital (HR 0.88 CI 0.49; 1.60, p=0.68). No plasma related serious adverse events were observed. The unadjusted ORs are available in the online supplement.

Altogether, the observations we made on antibody titers in patients and donors convinced us that a complete redesign of the study was needed and could not be resolved with a substantial study amendment. Indeed, we do not anticipate clinical benefit from ConvP for patients with high titers of autologous neutralizing antibodies present at baseline. The results were discussed with the DSMB on the 10^th^ of June and the decision was made to end the study under its current design.

## Discussion

The majority of the patients in the ConCOVID study already had high virus neutralizing antibodies titers on the day of study inclusion with titers comparable to the 115 recovered donors we screened for ConvP plasma donation. When the study was designed, the timing of neutralizing antibody development after SARS-CoV-2 infection was uncertain and we considered it unlikely that the most patients would have autologous neutralizing antibodies on the day of hospital admission. However, the data that became available to us after the inclusion of 86 patients made it very unlikely that the overall study population would benefit from ConvP therapy without a change in the patient recruitment strategy. Indeed, without a redesign of the study it would be substantially underpowered also after the enrollment of 426 patients and after a meeting with the DSMB the decision was made to end the study. No statistically significant differences in mortality (aOR 0.95, CI 0.20 – 4.67, p=0.95) or improvement in the day-15 disease severity (aOR 1.30, CI 0.52 - 3.32, p=0.58) was observed when the study was suspended.

We think that the observations we made are relevant for almost all ongoing studies on ConvP as a treatment for COVID-19. Indeed, all but few of these trials are focusing on hospitalized patients and the time from disease onset to admission was repeatedly shown to be comparable to the 10 days in our study. To the best of our knowledge, none of the ongoing studies is screening patients in real-time for the presence of antibodies before inclusion.^9,10^

In relation to the plasma donors that we selected, virtually all had anti-SARS-CoV-2 antibodies but only 41% had high neutralizing titers of at least 1:320. This could be related to generally milder disease course of the donors we recruited ^12,18^) Although, the level of antibodies required to ascertain an antiviral effect remains to be established a certain minimum level will be needed because after administration, the antibodies will be diluted at least 10-fold when administered to an adult patient. However, many of the ongoing trials are not directly testing the neutralizing capacity of donor plasma (which is the gold standard in coronavirus serology), but rather rely on a positive anti-SARS-CoV-2 ELISA or do not test for antibodies at all. Recent observations from our laboratory indicate that a positive Wantai SARS-CoV-2 Ig ELISA with an OD ratio ≥10 correlates well with a PRNT50 titer of at least 1:80.^17^ This could already help to reliably exclude donors with low neutralizing antibody when PRNT is unavailable, although it will not guarantee high PRNT50 titers. A solution may be to actively recruit recovered patients who had more severe COVID-19 disease as donor, as they have been shown to have higher antibody levels.^11,17^ Also, the use hyperimmune Ig preparations produced from pooled convalescent plasma (now called COVIg) or the use of specific highly neutralizing antibodies may solve this issue.^19,20^

This study has limitations. First, the premature ending of the study prevents definite conclusions regarding the clinical benefit of ConvP. Fortunately, a collaboration between several research groups evaluating ConvP is being formed and will allow for a pre-planned meta-analysis on pooled data from clinical trials. Our data do however show that ConvP should be studied earlier in the disease course. This could mean in the outpatient setting where ConvP could be evaluated in patients with a higher likelihood of disease progression based on clinical (e.g. age, comorbidities) or other (e.g. CRP, LDH) characteristics. Our data also show that in hospitalized patients testing for the presence of antibodies prior to ConvP should be part of the protocol, and stratifying or even excluding patients based on a positive antibody test will be needed. With the large variety of serological assays that have come available, it is important to carefully validate the assays prior to use and ideally correlate the assay to gold standard virus neutralization assays. Second, ConvP may have an effect that is unrelated to the neutralizing antibodies because therapy with intravenous immunoglobulines or plasma can have diverse anti-inflammatory and immunomodulatory effects as well. However, we considered any such effect unlikely because doses of Ig therapy when used as an immunomodulatory agent are typically at least 10-fold higher than the quantity of immunoglobulins in 300ml plasma. Finally, the recently completed dexamethasone arm of the recovery trial demonstrated an improved overall survival with dexamethasone therapy in patients requiring supplemental oxygen therapy.^1^ This will almost certainly change the standard of care therapy of COVID-19 and needs to be incorporated in the design of ongoing and future ConvP studies as well.

In conclusion, the majority of patients in the ConCOVID study already had high titers of virus neutralizing antibodies upon enrollment in the study. This made the design of the study unsuitable for its purpose of evaluating the clinical value of ConvP. This observation should trigger investigators to reconsider the design of current studies on ConvP for the treatment of patients with COVID-19.

## Data Availability

Anonymized data will be shared on reasonable request to non-for-profit organizations after review of the request and approval by the study team as long as the data sharing will serve a scientifically valuable purpose

## Acknowledgements

We would like to thank all plasma donors who volunteered in great numbers and all patients who participated in the study. All colleague internist-infectiologists for giving the study team at Erasmus MC the time to execute this study during unprecedented times. Prof. Stephanie Klein-Nagelvoort Schuit, prof. Annelies Verbon and prof. Charles Boucher for their support. The institutional review board at Erasmus MC for their efficient review of the application and several amendments on short notice. HOVON, in particular Monique Steijaert and Henk Hofwegen for helping with IRB related procedures and for programming the ALEA online randomization tool. Alders Lamore, for programming the eCRF. Sanquin Blood Supply Netherlands for collecting convalescent plasma from hundreds of donors. The members of the data safety monitoring board dr. JL Nouwen, prof. H Boersma, and dr. B Van der Hoven. The Erasmusfoundation for financial support and all those who donated to the foundation for this study. In particular Eduard Haegens and Marleen Hoex from Ypsilon who made it financially possible to start the trial. The department of virosciences and the department of immunology and all laboratory technicians and medical students (in particular Thijs Schrama, Renée Deckers, Freya Huijsmans, Niek van der Maas, Miliaan Zeelenberg, Zgjim Osmani, Jari Hofmans, Lieke Heijnen) for helping with real-time data and sample collections throughout the country. BMW, the Netherlands for providing two cars free of charge for sample collection. Dr. Corine Delsing from Medisch Spectrum Twente, dr. Jiri Wagenaar from Noordwest Ziekenhuisgroep and Robin Soetekouw from Spaarne Gasthuis for engaging as a study site. All study nurses and trial coordinators in the participating centres, and in particular Siepke Hiddema, Frances Greven and resident Sander Albers, and many others that volunteered their time and effort for this study.

## Appendix

The authors’ full names and academic degrees are as follows: Arvind Gharbharan, M.D. Carlijn C.E. Jordans, M.D, Corine Geurtsvankessel, M.D., PhD, Jan G. den Hollander, M.D., PhD, Faiz Karim, M.D., PhD, Femke P. N. Mollema, M.D., PhD, Janneke E. Stalenhoef – Schukken, M.D., PhD, Anthonius Dofferhoff, M.D., PhD, Inge Ludwig, M.D., Adrianus Koster, M.D., Robert-Jan Hassing, M.D., PhD, Jeanette C. Bos, M.D, PhD, Geert R.van Pottelberge, M.D., PhD, Imro N. Vlasveld, M.D., Heidi S. M. Ammerlaan, M.D., PhD, Elena M. van Leeuwen – Segarceanu, M.D., PhD, Jelle Miedema, M.D., Menno M. van der Eerden, M.D., PhD, Grigorios Papageorgiou, Msc., Peter A.W. te Boekhorst, M.D., PhD, Francis H. Swaneveld, M.D., Peter D. Katsikis, M.D., PhD, Yvonne Mueller, PhD, Nisreen M.A. Okba, PhD, Marion P.G. Koopmans, PhD, ., Bart L. Haagmans, PhD, Casper Rokx, M.D., PhD, Bart J.A. Rijnders, M.D., PhD.

The authors’ affiliations are as follows: Erasmus MC, University Medical Center, Rotterdam (A.G., C.J., C.G., J.M., M.E., G.P., P.B., P.K., Y.M., N.O., M.K., B.H., C.R., B.R.), Maasstad Hospital, Rotterdam (J.H.), Groene Hart Hospital, Gouda (F.K.), Haaglanden Medical Center, the Hague (F.M.), Onze Lieve Vrouwe Hospital, Amsterdam (J.S.), Canisius Wilhelmina Hospital, Nijmegen (A.D.), Bernhoven Hospital, Uden (I.L), VieCurie Medical Center, Venlo (A.K.), Rijnstate Hospital, Arnhem (R.H), Reinier de Graaf Hospital, Delft (J.B.), ZorgSaam Hospital, Terneuzen (G.P.), Martini Hospital, Groningen (I.V.), Catharina Hospital, Eindhoven (H.A.), Sint Antonius Hospital, Nieuwegein (E.L.), Unit of Transfusion Medicine, Sanquin Blood Supply, Amsterdam (F.S)

All in the Netherlands

## Appendix 2

Full protocol

## Appendix 3

Additional methods and results

